# Prediction of BAP1 mutations in uveal melanoma patients from histology images using weakly supervised deep learning-based whole slide image analysis

**DOI:** 10.1101/2021.09.16.21263694

**Authors:** Garv Mehdiratta

## Abstract

While cases of uveal melanoma are relatively rare overall, it remains the most common intraocular cancer in adults and has a 10-year fatality rate of approximately 50% in metastatic patients with no effective treatment options. Mutations in BAP1, a tumor suppressor gene, have been previously found to be associated with the onset of metastasis in uveal melanoma patients. In this study, I utilize a weakly supervised deep learning-based pipeline in order to analyze whole slide images (WSIs) of uveal melanoma patients in conjunction with slide-level labels regarding the presence of BAP1 mutations. I demonstrate that there is a strong relationship between BAP1 mutations and physical tumor development in uveal melanoma and that my model is able to predict such relationships with an optimized mean test AUC of 0.86. My findings demonstrate that deep learning models are able to accurately predict patient-specific genotypic characteristics in uveal melanoma. Once integrated into and adapted to existing non-invasive ocular scanner technologies, my model would assist healthcare professionals in understanding the specific genetic profiles of their patients and provide more personalized treatment options in a safe, efficient manner, thus resulting in improved treatment outcomes.

## Introduction

Uveal melanoma is a type of eye cancer involving the uveal tract, or middle layer, of the eye. The uvea consists of three parts: the iris, the ciliary body, and the choroid. Collectively, these structures are responsible for a variety of ocular functions, including controlling the amount of light that enters the eye and providing nutrition to the retina. Uveal melanoma is caused by tumors forming in melanocytes, which are responsible for producing melanin for eye pigmentation, and although cancer can originate from melanocytes in any of the three regions of the uveal tract, most cases involve the choroid [1]. For instance, in a study analyzing the tumor characteristics of 8,033 patients, 90% of cases had tumors that originated in the choroid [2]. Throughout this section, I will discuss my own analysis of a uveal melanoma patient dataset obtained from The Cancer Genome Atlas, and I will explore a more in-depth review of this dataset later in this report. Within this dataset, I discovered that 83.7% out of the 80 total patients had tumors that originated in the choroid as depicted in Fig 1, affirming that my data was not significantly skewed regarding the tumor region of origin using the results of the earlier study as a baseline for comparison.

**Fig 1.**
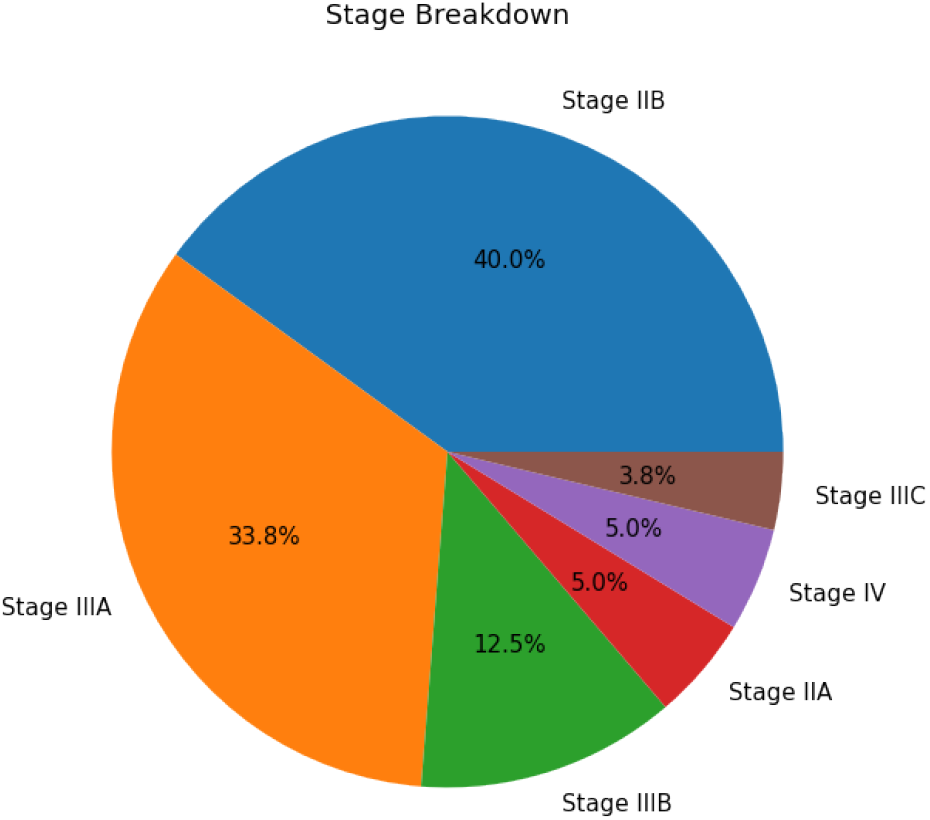
Breakdown of cancer stage for uveal melanoma patients in the 80-patient TCGA-UVM dataset. Most patients had intermediate-stage cancer, with 40.0% of patients with Stage IIB cancer and 33.8% of patients with Stage IIIA cancer. Patients with stage IIC cancer were least common, at just 3.8%.

Uveal melanoma is the most prevalent eye-related disease [1]. However, incidence rates for the disease are relatively low and as a result, there is a lack of studies regarding uveal melanoma. In the United States, for instance, an analysis of the National Cancer Institute’s Surveillance, Epidemiology, and End Results (SEER) program database reveals that from 1973 to 2008, the age-adjusted overall mean incidence of uveal melanoma remained constant at around 5.1 new cases per million people [3]. Throughout this period, survival rates did not improve even as more conservative treatments were employed, demonstrating the lack of effective treatments available to uveal melanoma patients. Interestingly, incidence rates have been found to be associated with a patient’s geographic location; an analysis of data obtained from the European Cancer Registry indicates that a north-to-south decreasing gradient in uveal melanoma diagnoses is present [4]. This disparity is due to a protective effect imparted by ocular pigmentation on people living in lower latitudes due to their relatively high level of exposure to ultraviolet light as compared to those living in higher latitudes [4].

Past studies have found that the elderly have higher incidence rates of uveal melanoma, with peak occurrences at around 70 years of age [3]. This conclusion is supported by my own analysis of the TCGA-UVM cohort, as patients in the 70-79 age bracket comprise 22% of the total number of cases in the dataset, which is the highest percentage among all the age groups. A graph of this distribution resembles a skewed normal curve, with about 60% of the caseload belonging to the 50-79 age group (as shown in Fig 2). Previous population-based studies have also shown that generally, males tend to have higher incidence rates than females [4, 5]. This trend is also supported by my analysis of the TCGA dataset, as 56.2% of the uveal melanoma cases were of men, while only 43.8% belonged to women (Fig 3. Race also has a strong relationship with incidence rates: in a study of 8100 uveal melanoma patients, 98% of them were Caucasian [6]. While researchers are unclear as to exactly why this is, potential explanations include the impact of darker skin pigmentation and other cultural, environmental, and socioeconomic factors prevalent in non-Caucasians as well as implicit bias against non-Caucasians.

**Fig 2.**
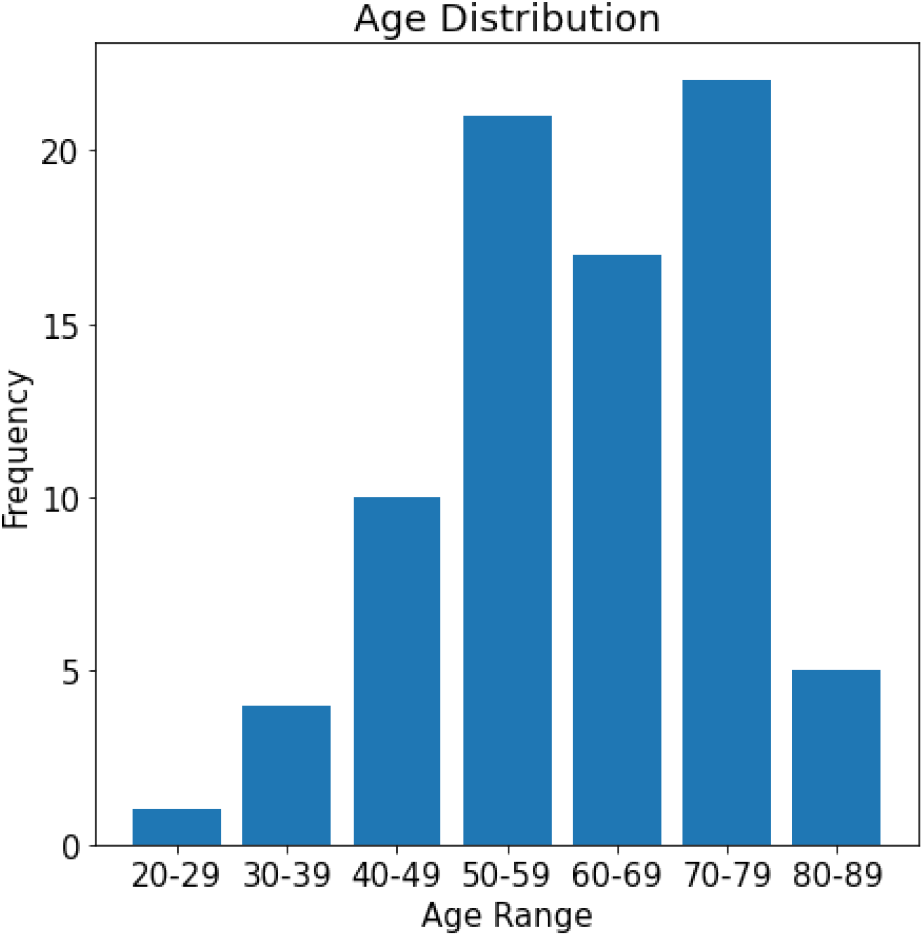
Age distribution for the studied patients in the 80-patient TCGA-UVM dataset. Over 50% of the uveal melanoma patients were between 50 and 79 years of age. The graph resembles a negatively skewed normal curve, with the center of the distribution shifted to the right from the center of the graph (at the label corresponding to 50-59 years of age).

**Fig 3.**
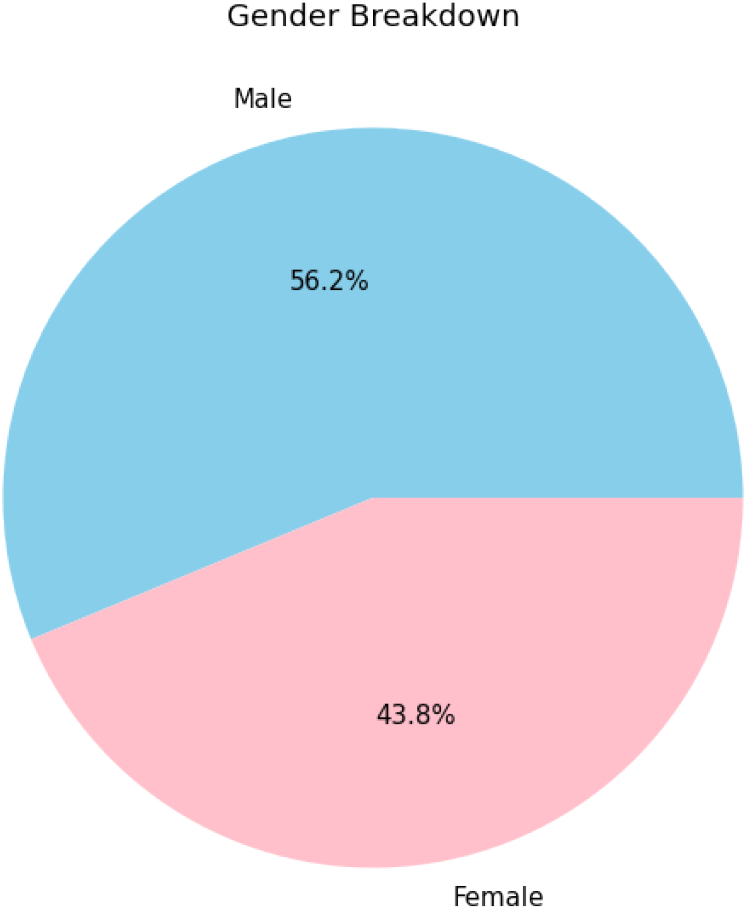
Breakdown of patient gender in the 80-patient TCGA-UVM dataset. There was a slightly higher percentage of males as compared to females in the patient dataset: 56.2% of patients in the dataset were male, while 43.8% were female.

There are multiple phenotypic characteristics that have been found to be risk factors for uveal melanoma. These characteristics include fair/light skin tone, freckles, blond hair, light eye color, and an inability to tan, all features that are more commonly found in Caucasian individuals [7]. As such, it is evident that Caucasians generally have a higher risk of experiencing uveal melanoma than non-Caucasians are, supporting the previously mentioned trend of a large majority of uveal melanoma patients being Caucasian.

Additionally, there are also some genotypic characteristics that are associated with the development and progression of uveal melanoma. One gene that has been identified as playing a role in the progression of uveal melanoma is the BAP1 (BRCA1-associated protein 1) gene. Located at Chromosome 3p21.1, BAP1 is a tumor suppressor gene that encodes a nuclear ubiquitin carboxy-terminal hydrolase, a type of deubiquitinating enzyme [8]. This allows the gene to regulate multiple cell division-related processes, including DNA damage repair, cell cycle control, and programmed cell death. Previous clinical studies have found that mutations in the BAP1 gene are accurate predictors of uveal melanoma metastasis in patients [9]. For example, a study found that patients who had a concurrent BAP1 mutation that led to a negative BAP1 gene expression were 7.7 times more likely to develop uveal melanoma metastases than patients who did not have the mutation [10]. As such, due to its significance in the progression of uveal melanoma, I will be analyzing the BAP1 gene and its relationship to tumor development as depicted in whole slide images (WSIs) of uveal melanoma patients in this project.

## Materials and methods

### Data

The data for this project was obtained from The Cancer Genome Atlas (TCGA), a program developed by the National Cancer Institute and the National Human Genome Research Institute that offers a wide array of clinical and biomedical data for 33 types of cancer free of charge to the general public. For this project, I utilized 80 diagnostic histology slides of uveal melanoma patients and genetic mutation data for each patient from the TCGA Genomic Data Commons Portal, which I used to train my model. Within the TCGA-UVM cohort, 25 out of the 80 total patients were found to have mutations in the BAP1 gene, exhibiting a skew towards non-mutation cases.

### Model

To analyze the slides of uveal melanoma patients obtained from the TCGA program, I leveraged CLAM, a weakly-supervised deep learning-based pipeline for analysis of whole slide images. CLAM uses attention-based learning and clustering in order to identify WSI subregions of high diagnostic value with regard to slide labels [11]. This pipeline consists of 3 main steps: WSI segmentation and patching, training using slide-level labels, and WSI heatmap construction using regions of interest (ROIs) identified by the model [11]. These attention maps are able to accurately identify the areas of highest importance and relevance in disease development and progression [11].

Instead of using the relatively inflexible max-pooling aggregation function, the main model uses the attention-based pooling function, which is trainable and allows for multi-class classification [11]. The first fully connected layer of the model serves to compress fixed 1024-dimensional patch-level representations to a 512-dimensional vector. Additionally, the model includes a binary clustering layer with 512 hidden units after the first fully connected layer. This instance-level clustering layer assigns pseudo labels to individual patches due to the lack of patch-level labels and optimizes the patches that the model strongly attends to or ignores, thus ensuring that strong positive evidence for each class is separable from strong negative evidence and facilitating the ability of the model to learn on class-specific features [11]. Throughout the training process, the model calculates “attention scores” for each WSI patch. These attention scores represent how important a particular patch is to the final slide-level classification result and reflect the ability of the model to perform a weakly supervised, multiple instance learning-based analysis of the data [11]. For example: for a given slide, if the overall slide-level label is classified as *L*, patches with a high attention score will be interpreted as being strong positive evidence for class *L*, while patches with a low attention score will be interpreted as being strong negative evidence for class *L*.

### Preprocessing and training

I began the data preprocessing stage by segmenting the tissue regions of the slides and separating them into individual 256 × 256-pixel patches. A ResNet-50 CNN model pre-trained on the ImageNet dataset was then used to extract features from the patches in the form of 1024-dimensional feature embeddings. Training the model on these features instead of the individual slide images resulted in increased efficiency, because the conversion of WSIs to feature vectors vastly decreased the amount of required data space and thus also decreased the amount of computational power needed to train the model. Prior to training, I queried the TCGA GDC data portal for the genetic profiles of each uveal melanoma patient and constructed a label CSV file consisting of binary values representing the presence of BAP1 mutations in the 80 patients. This file was used by the main multiple instance learning-structured model (obtained from the CLAM pipeline) during training in conjunction with the WSI features that were created in the preprocessing stage in order to determine correlations and generate predictions based on the provided feature and label data.

When running the model on the feature dataset, I utilized 5-fold monte carlo cross-validation, and for each fold, I allocated 80% of the dataset for training, 10% for validation, and the remaining 10% for testing. These splits were generated randomly and I ensured the accuracy of my results by running the model multiple times using different training data splits for each iteration.

During this project, my primary programming language was Python (version 3.7.11), and I implemented and ran all code in a Google Colaboratory notebook environment using an NVIDIA Tesla P100 GPU.

## Results

On the TCGA-UVM dataset with BAP1 slide-level binary labels and a learning rate of 0.0001, I achieved a 5-fold mean test AUROC of 0.86 (Fig 4). Based on the calculated J Statistic, I implemented an optimal threshold of 0.3306 during mutation vs. non-mutation classification, the results of which are shown in Table 1. This threshold was used to account for the uneven nature of the mutation data since around 68% of patients in the dataset did not have BAP1 mutations, creating an imbalance.

**Table 1.** Classification summary report for predictions based on the optimal cutoff value of 0.3306.

| Classification Summary Report |  |  |  |
| --- | --- | --- | --- |
|  | Precision | Recall | F1-Score |
| BAP1-Negative | 1.0000 | 0.8000 | 0.8889 |
| BAP1-Positive | 0.6667 | 1.0000 | 0.8000 |
| Accuracy | 0.8571 | 0.8571 | 0.8571 |
| Macro Avg. | 0.8333 | 0.9000 | 0.8444 |
| Weighted Avg. | 0.9048 | 0.8571 | 0.8635 |

**Fig 4.**
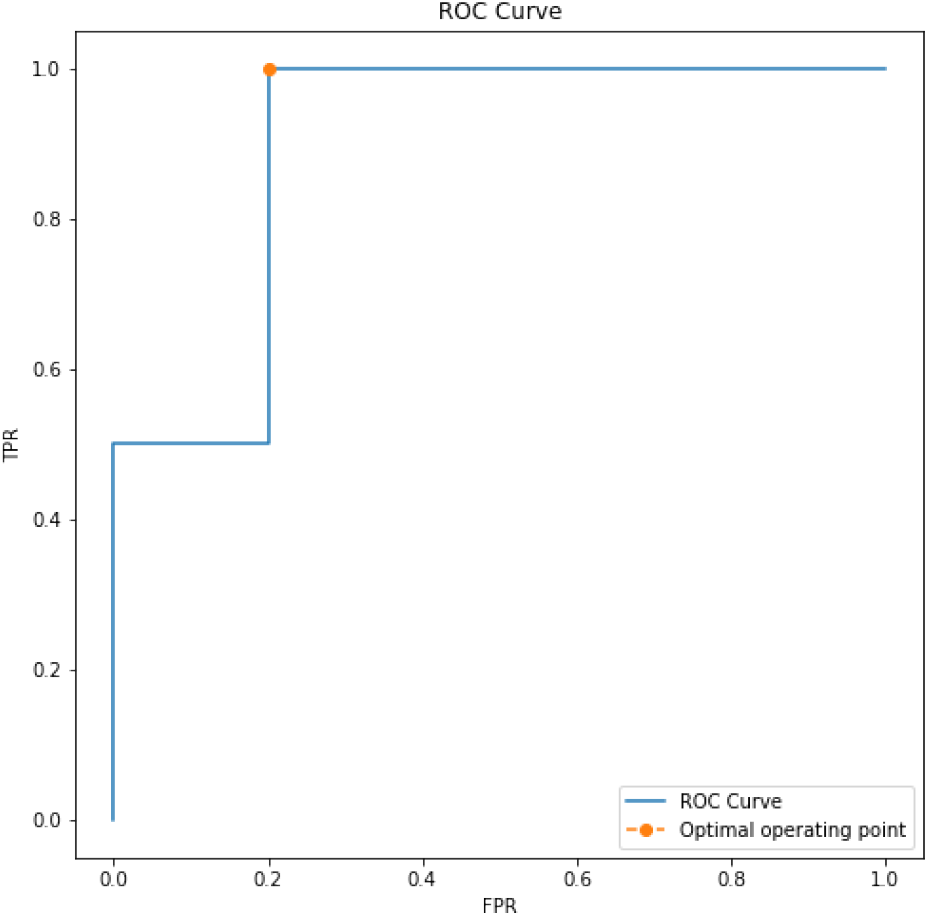
ROC curve representing the maximum AUC value achieved by the model. The mean AUC score across all training/testing splits was 0.86.

Based on my analysis of available studies, BAP1 mutations have been previously linked to metastasis but not physical tumor development. My results demonstrate that within the available data, there is a strong relationship between BAP1 mutations and physical tumor development. The results also demonstrate that deep learning models are able to accurately predict patient-specific genotypic characteristics in uveal melanoma. As such, since BAP1 mutations serve as a predictor of cancer metastasis, these conclusions are very significant because they imply that deep learning-based technologies could be used to identify the risk of uveal melanoma metastasis in patients simply based on the presence of BAP1 mutations.

## Discussion

Because up to 50% of uveal melanoma patients who experience metastasis die within 10 years, it is of the utmost importance for medical professionals to be able to accurately predict whether their patients will likely experience metastasis in a non-invasive manner in order for the most effective patient-based treatment plans to be developed and implemented. While such approaches are not universally possible in the present day, future technologies could be developed using a deep learning prediction tool such as the one developed in this project.

In fact, such tools are already in development for other eye-related diseases; for example, researchers at Google Health are developing a novel AI-based tool that is able to non-invasively perform an eye scan and predict whether signs of diabetic retinopathy are present with 90% accuracy in a process that lasts less than 10 minutes [12]. If applied to uveal melanoma, oncologists would be able to discover the presence of BAP1 mutations in their patients in a non-invasive, rapid manner, maximizing the amount of potential treatment opportunities while minimizing harm to the patient. Such technological innovations would be especially useful in low-resource settings where hospitals and health clinics may lack the necessary resources (specialized medical personnel, medical equipment, etc.) to be able to collect and analyze histopathological images from patients to aid in disease detection.

The BAP1 gene should also be explored as a potential therapeutic target for future treatments being developed for uveal melanoma. If a drug is developed that allows patients with BAP1 mutations whose tumors are still localized to regain BAP1 gene function, future metastasis could be avoided, promoting the likelihood of favorable treatment outcomes. Such a drug would likely be most effective if given to BAP1-negative patients before metastasis occurs, emphasizing the importance of non-invasive deep learning-based tools that allow for the detection of BAP1 mutations in patients as early as possible, ultimately minimizing the need for more invasive methods like enucleation in order to analyze physical ocular tissue and conduct a pathology analysis.

One major limitation of this study was the relatively small dataset of 80 uveal melanoma WSIs I had access to through the TCGA program. With a larger dataset, my model would be able to analyze more obscure intricacies of WSI features and make much more accurate predictions regarding the presence of BAP1 mutations in patients, minimizing the false positive rate and maximizing the true positive rate as much as possible. While my AUC score of 0.86 is promising, it could be improved upon by fine-tuning the model by training on more images, thereby improving the accuracy of the model’s classification ability.

Although my analysis was focused on the relationship between BAP1 genetic mutations and uveal melanoma WSI features, I acknowledge and plan to explore the numerous available opportunities to expand my findings from this project. Engaging in a computational pathology-based study of the impact of BAP1 mutations across all 33 cancers in the TCGA program, for example, would allow me to discover whether such mutations affect other cancers in a similar manner to their effects on uveal melanoma progression. Within the scope of uveal melanoma, exploring other commonly mutated genes as I did with BAP1 could lead to an enhanced understanding of how other genetic mutations affect physical tumor development. Both of these approaches ultimately serve to open the door to and lay the foundation for rapid, non-invasive, mutation-based cancer detection as well as targeted therapy and treatment development.

## Data Availability

All uveal melanoma WSI data was obtained from The Cancer Genome Atlas, a program developed by the National Cancer Institute and the National Human Genome Research Institute that offers a wide array of clinical and biomedical data for 33 types of cancer free of charge to the general public.

https://portal.gdc.cancer.gov/

## Supporting information

**S1 Fig.**
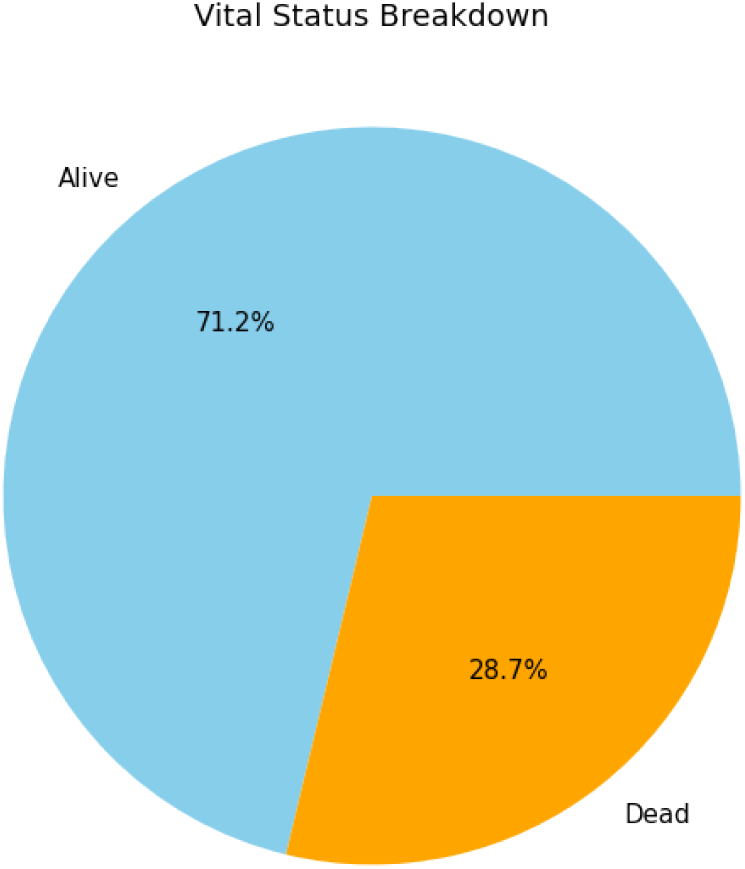
Breakdown of vital status for the 80 uveal melanoma patients in the 80-patient TCGA-UVM dataset. At the time of data collection, 71.2% of uveal melanoma patients were alive, while the remaining 28.7% were dead.

**S2 Fig.**
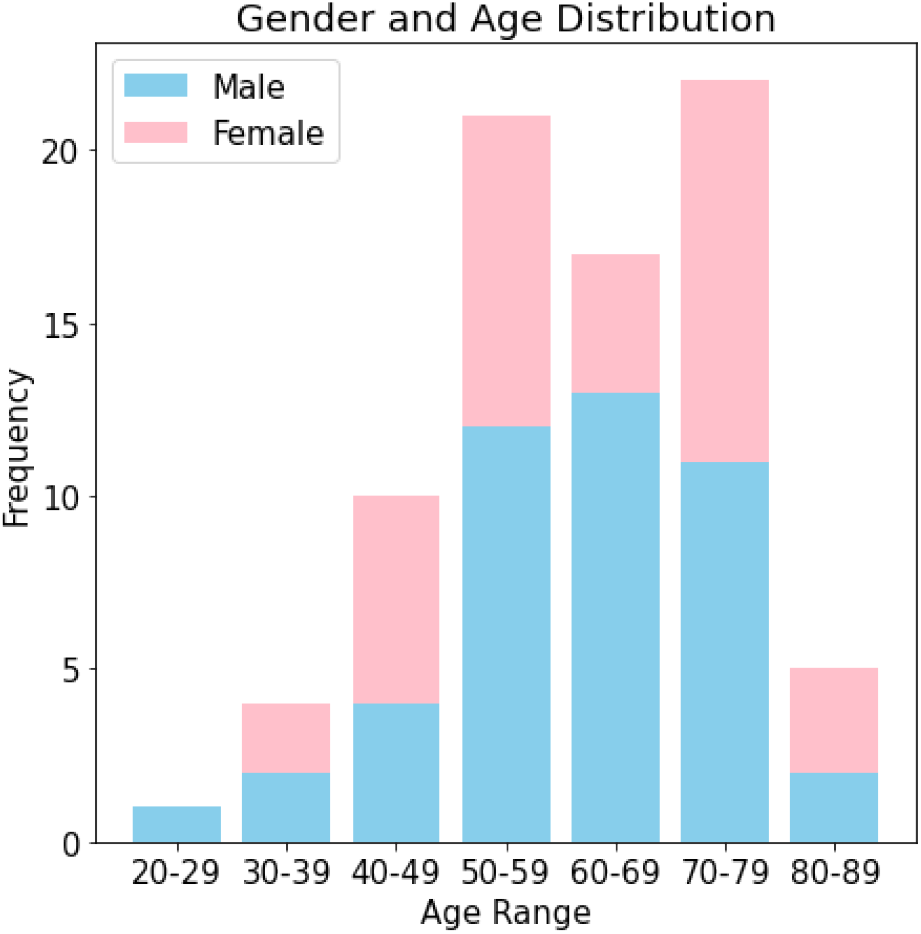
Gender distribution for each patient age range represented in the 80-patient TCGA-UVM dataset. Taking into account the presence of slightly more male patients than female patients in the dataset, both genders were relatively evenly distributed across all represented age ranges.

**S3 Fig.**
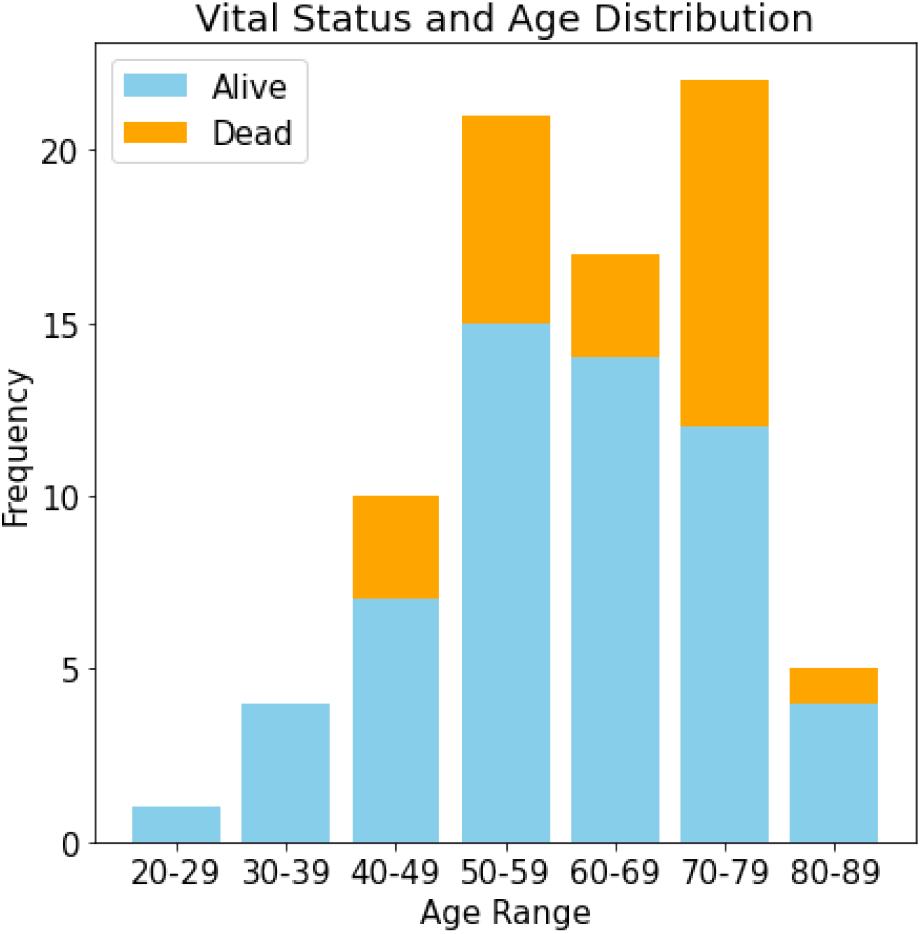
Vital status distribution for each patient age range represented in the 80-patient TCGA-UVM dataset. The fatality rate for younger patients seemed to be relatively lower than that of older patients, which could be a result of more advanced cancer progression in elderly patients who have had uveal melanoma for a greater length of time than younger patients. However, further research is needed to verify this trend.

**S4 Fig.**
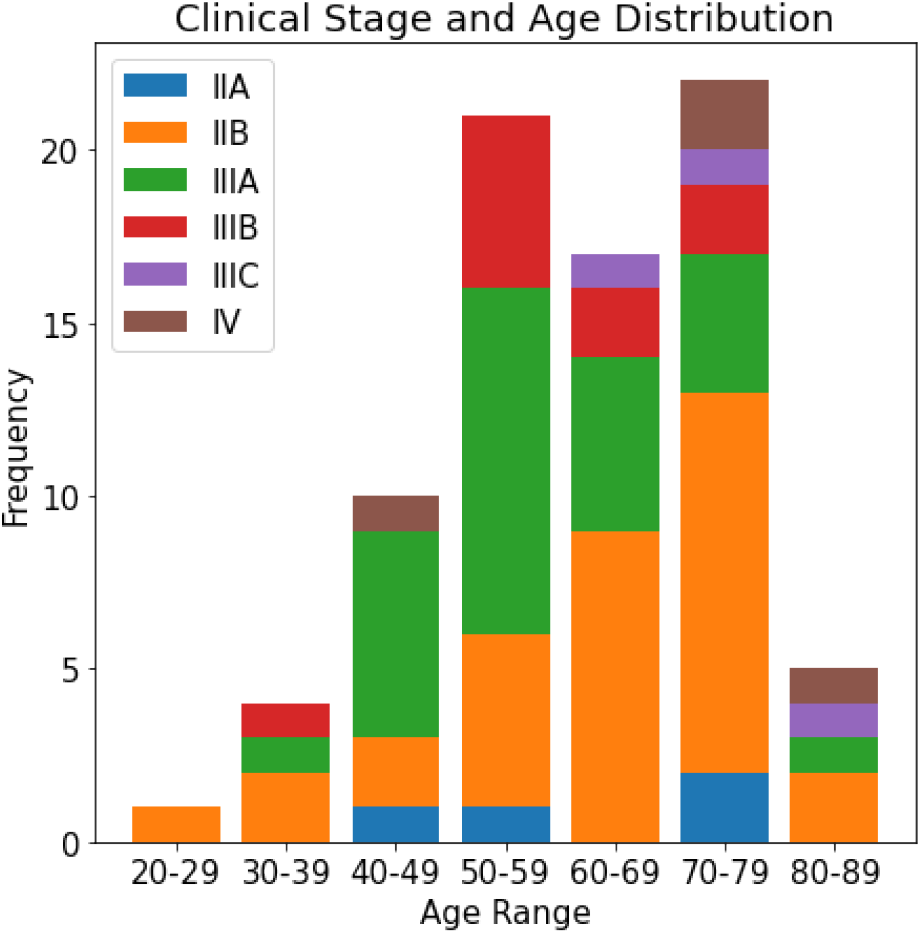
Cancer clinical stage distribution for each patient age range represented in the 80-patient TCGA-UVM dataset. Stage IIB and IIIA are the most prevalent cancer stages among the 80 patients, and more advanced stages seem to be more prevalent in more elderly patients.

**S5 Fig.**
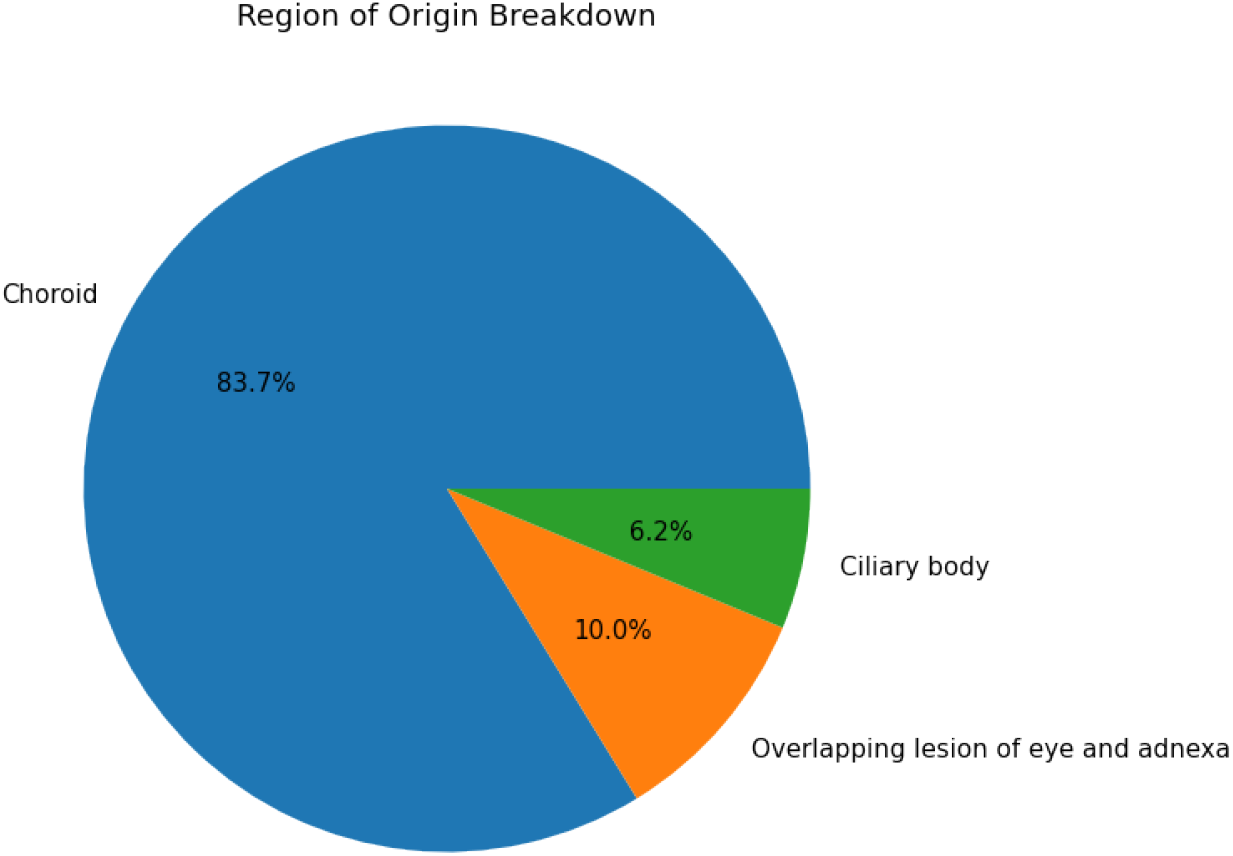
Cancer region of origin distribution for each patient age range represented in the 80-patient TCGA-UVM dataset. For all age ranges, the large majority of patients have tumors that originated in the choroid, with the other two possible regions of origin (ciliary body and OLEA/overlapping lesion of eye and adnexa) relatively evenly distributed across all age ranges.

## Acknowledgments

The author would like to thank Sharifa Sahai, Department of Systems, Synthetic, and Quantitative Biology at Harvard Medical School, for her feedback on the manuscript and invaluable guidance throughout the duration of this project.

